# Development of a Machine Learning Model for Predicting In-Hospital Mortality and Analyzing Associated Risk Factors Using Large Patient Samples

**DOI:** 10.1101/2025.02.27.25323062

**Authors:** Jinxin Liu, Haoyue He, Yanglingxi Wang, Jun Du, Kaixin Liang, Jun Xue, Yidan Liang, Peng Chen, Qiang Yang, Ying Yin, Guixue Wang, Xue Jiang, Yongbing Deng

**Author notes:** Corresponding author: Yongbing Deng Xue Jiang. These authors have contributed equally to this work and share first authorship (Jinxin Liu, Haoyue He).

## Abstract

**Objective:** This study endeavors to construct a machine learning model to forecast in-hospital mortality and dissect associated risk factors, utilizing a vast dataset from multiple hospitals in Chongqing.

**Methods:** We amassed detailed baseline data encompassing demographics, medical histories, laboratory tests, and imaging indicators from 23,307 ischemic stroke patients. The NIHSS score was derived from admission records, and both in-hospital survival status and causes of death were meticulously documented. Employing the missForest method, we imputed missing values, addressing data imbalance through random oversampling, validated via five-fold cross-validation. The SHAPRFECV technique was instrumental in identifying the most impactful features, steering clear of multicollinearity. A suite of machine learning models, including LR, RF, and KNN, were meticulously tuned using three-fold cross-validation and grid search to optimize hyperparameters.

**Results:** Our cohort had an average age of 67.347 ± 12.822 years, a baseline NIHSS score of 8.430 ± 3.162, and a 51.186% male predominance, with an in-hospital mortality rate of 6.183%. The Random Forest model excelled with an AUC of 0.940 in the test set, trailed closely by CatBoost at 0.937, LightGBM at 0.930, and XGBoost at 0.929. Notably, CatBoost boasted the highest F1 score of 0.595420 on the test set, with no significant predictive performance disparity between it and the Random Forest model (p = 0.500).

**Conclusion:** Grounded in data from four hospitals in Chongqing, our machine learning model, predicated on baseline features, not only streamlines clinical application but also ensures robust predictive efficacy. It provides an in-depth analysis of mortality risk factors, serving as a pivotal reference for clinical decision-making. Future endeavors will concentrate on validating the model within larger-scale, geographically diverse samples, thereby amplifying its applicability and value in clinical practice.

## Introduction

### Background and Mortality of Ischemic Stroke

Stroke is a leading cause of death and disability worldwide. Statistics indicate that in 2021, 3.71 million people died from ischemic stroke globally, and it is projected that by 2030, the number of ischemic stroke cases will rise to 9.62 million, resulting in 2.45 million deaths. In China, the situation regarding cardiovascular diseases is even more severe, with the number of deaths due to stroke increasing by 171.0% from 1990 to 2019, a trend that continues to persist. Approximately 80% of stroke cases are ischemic, primarily caused by arterial occlusion, leading to local brain tissue ischemia and hypoxia, ultimately resulting in brain cell necrosis. Therefore, accurately predicting the prognosis of ischemic stroke patients, especially through early individualized assessments at the time of admission, is crucial. This will assist clinicians in timely implementing appropriate treatment measures, thereby improving patient outcomes.

### Previous Research

Various factors have been identified that can predict and influence patient prognosis, including diabetes, prior heart disease, high sodium diets, high cholesterol, and baseline CRP levels. However, no single biomarker has been found to achieve accurate predictions, and relying on a single biomarker may not adequately reflect the underlying complexities. Increasingly, research has begun to comprehensively integrate clinical data from patients to establish multi-biomarker predictive models.

Multiple predictive scoring systems have been developed for the mortality of ischemic stroke patients, including the NIHSS score, iSCORE, SOAR, and GWTG-Stroke scores. Among these, Saumya et al. reported the best-performing model, which they compared to other stroke prognosis scores (such as ASTRAL, DRAGON, FSV, iSCORE, PLAN, and THRIVE), finding that their model’s AUC for predicting various functional and mortality outcomes at 3-12 months post-stroke ranged from 0.71 to 0.89. However, many currently available predictive models are based on data collected years ago or use data over a longer time span to increase sample size, and none have been widely implemented in clinical practice. Additionally, given the significant changes in stroke care over the past 20 years, this may hinder the effective implementation of these models in current clinical routines. Overall, the quality of stroke prognosis models varies widely, and existing models for predicting post-stroke mortality are limited by sample size, breadth of clinical variables, and overall clinical utility, lacking a reasonable and clinically applicable model.

### Machine Learning

In recent years, machine learning algorithms have gained popularity, demonstrating significant advantages over traditional methods in identifying and screening important risk factors. These algorithms excel at handling large and complex clinical datasets, overcoming the limitations of traditional linear scoring models, making them more suitable for addressing complex nonlinear data associations. However, common issues exist in the data used in routine clinical practice, such as multicollinearity, missing values, and outliers. Additionally, smaller sample sizes may lead to overfitting or underfitting of models, resulting in overly optimistic or even erroneous outcomes. The opacity of machine learning models may also diminish clinicians’ confidence in their application. Furthermore, some machine learning models incorporate too many predictive variables, reducing their practicality in emergency situations. Overall, the application of new technologies such as machine learning offers new possibilities for risk prediction and management in stroke patients. These technologies not only enhance the accuracy of patient prognosis assessments but also assist clinicians in formulating personalized treatment plans, thereby improving patients’ quality of life. With ongoing research into the causes and mechanisms of ischemic stroke, more effective interventions and treatments are expected to be developed, positively impacting the reduction of stroke incidence and improving patient outcomes.

This study aims to utilize large sample patient data from multiple hospitals in Chongqing, broadly incorporating historical features, laboratory test data, and imaging indicators. By increasing data quantity and dimensionality and conducting interpretability analyses, this research seeks to overcome the limitations of machine learning as much as possible. The goal is to develop a machine learning model that predicts in-hospital mortality based on baseline features, maximizing model simplicity to enhance its convenience in clinical use while ensuring predictive efficacy and analyzing related risk factors.

## Results

### Patient Characteristics

In the study, a total of 23,307 patients diagnosed with ischemic stroke were analyzed, as shown in the inclusion and exclusion flowchart in Figure 1. The demographic and clinical characteristics of these patients are summarized as follows: the average age was 67.347 years, with a standard deviation of 12.822 years, indicating a relatively broad age distribution. The mean baseline National Institutes of Health Stroke Scale (NIHSS) score was 8.430, with a standard deviation of 3.162, reflecting the severity of stroke symptoms at admission. The study population included 51.186% males, highlighting a slight male predominance. Additionally, the overall in-hospital mortality rate was recorded at 6.183%, which is an important metric for evaluating the prognosis of patients with ischemic stroke.

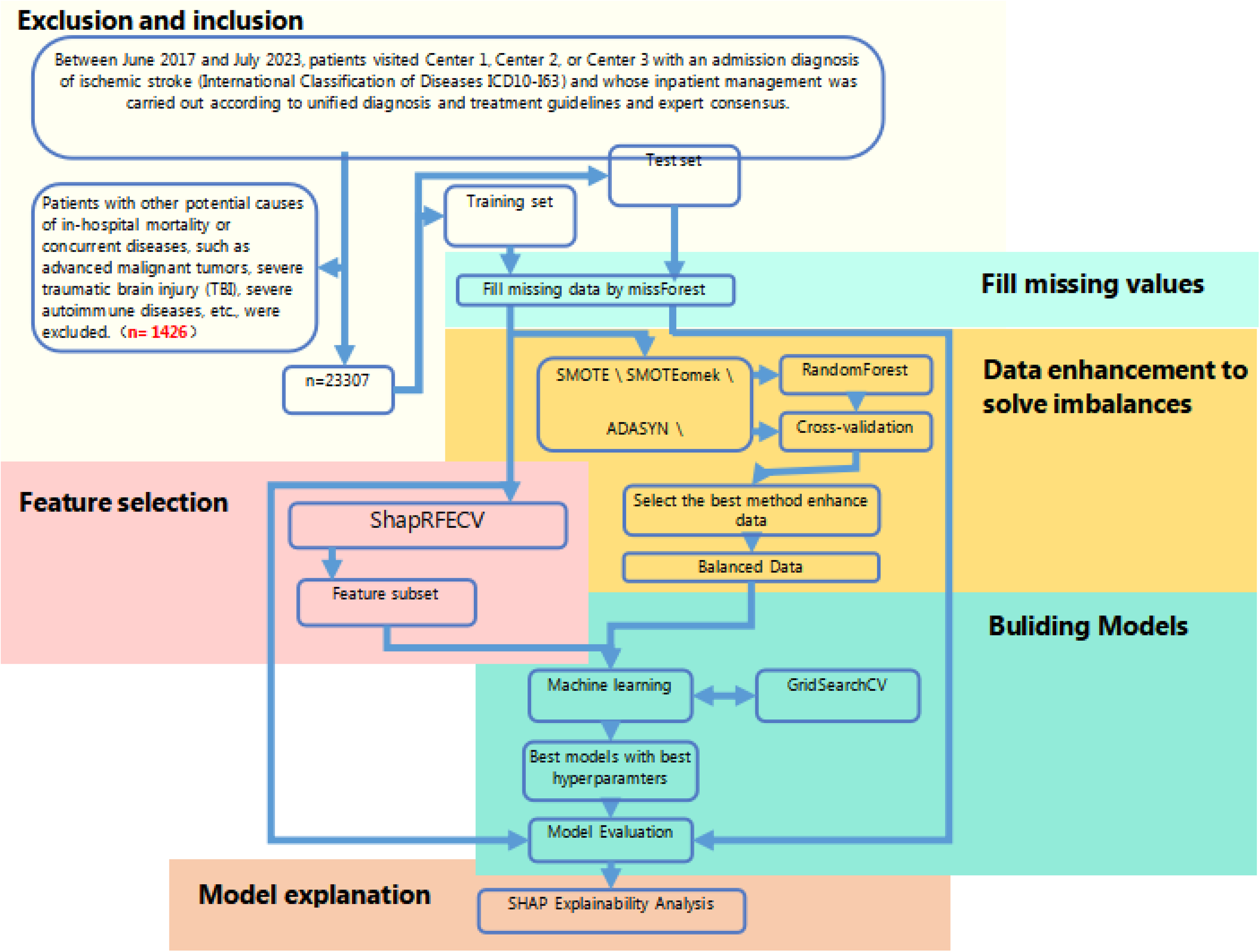

To further investigate the predictive factors associated with in-hospital mortality, univariable and multivariable analyses were conducted. The results of these analyses are presented in detail in Table 1 and Table 2.

After comparing several data balancing methods, random oversampling yielded the best results, and this method was employed for data balancing, as shown in Figure 2.

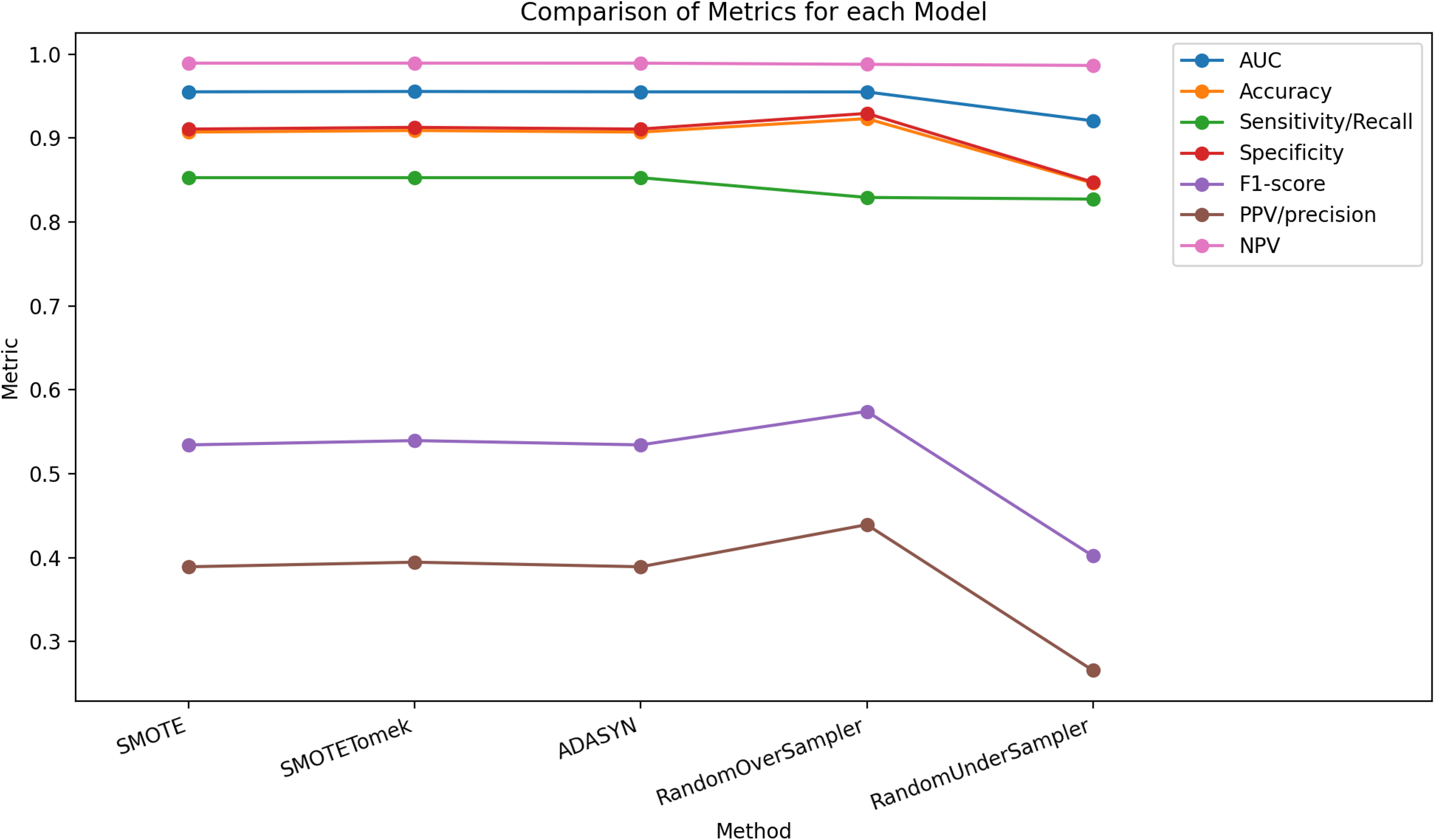

### Feature Selection

The SHAPRFECV feature selection process is illustrated in Figure 3. When the number of selected features was limited to 13 (Feature Subset A), the predictive performance of the model was comparable to that using a larger number of features. The 13 selected features included: creatinine, NIHSS score, ischemia-modified albumin, blood urea nitrogen, gender, uric acid, D-dimer, baseline brain herniation, C-reactive protein, chloride levels, aspartate aminotransferase, anion gap, and potassium levels. When restricting the number of features to 6, the selected features (Feature Subset B) were D-dimer, NIHSS score, C-reactive protein, blood urea nitrogen, gender, and creatinine.

Model Performance

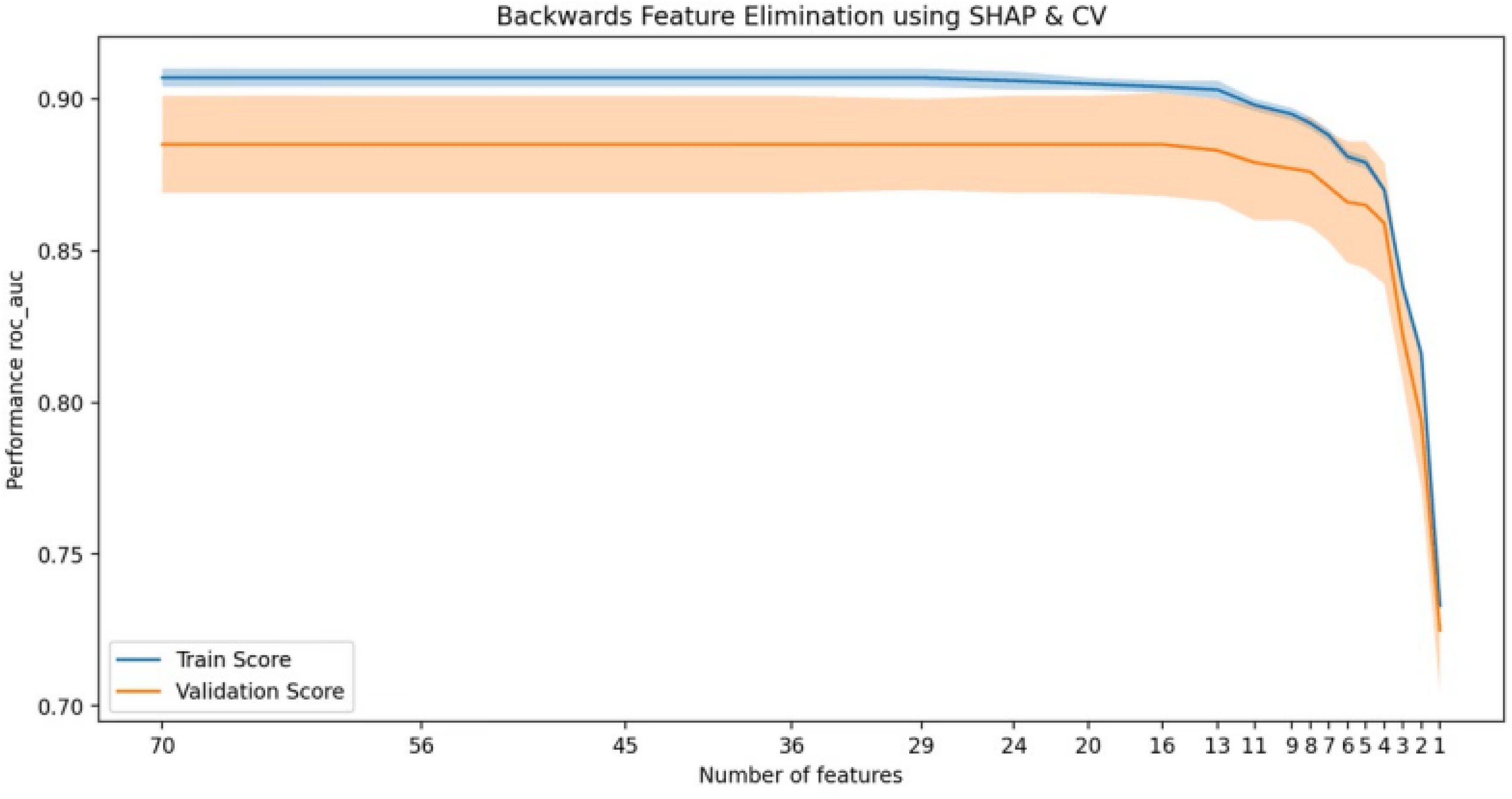

### Full Model

Figure 4 displays the ROC curves for various predictive models constructed using Feature Subset A (13 features). Table 4 includes model performance evaluation results of Full Model. The Random Forest (RF) model achieved the highest AUC of 0.940 on the test set, followed by CatBoost at 0.937, LightGBM at 0.930, and XGBoost at 0.929.

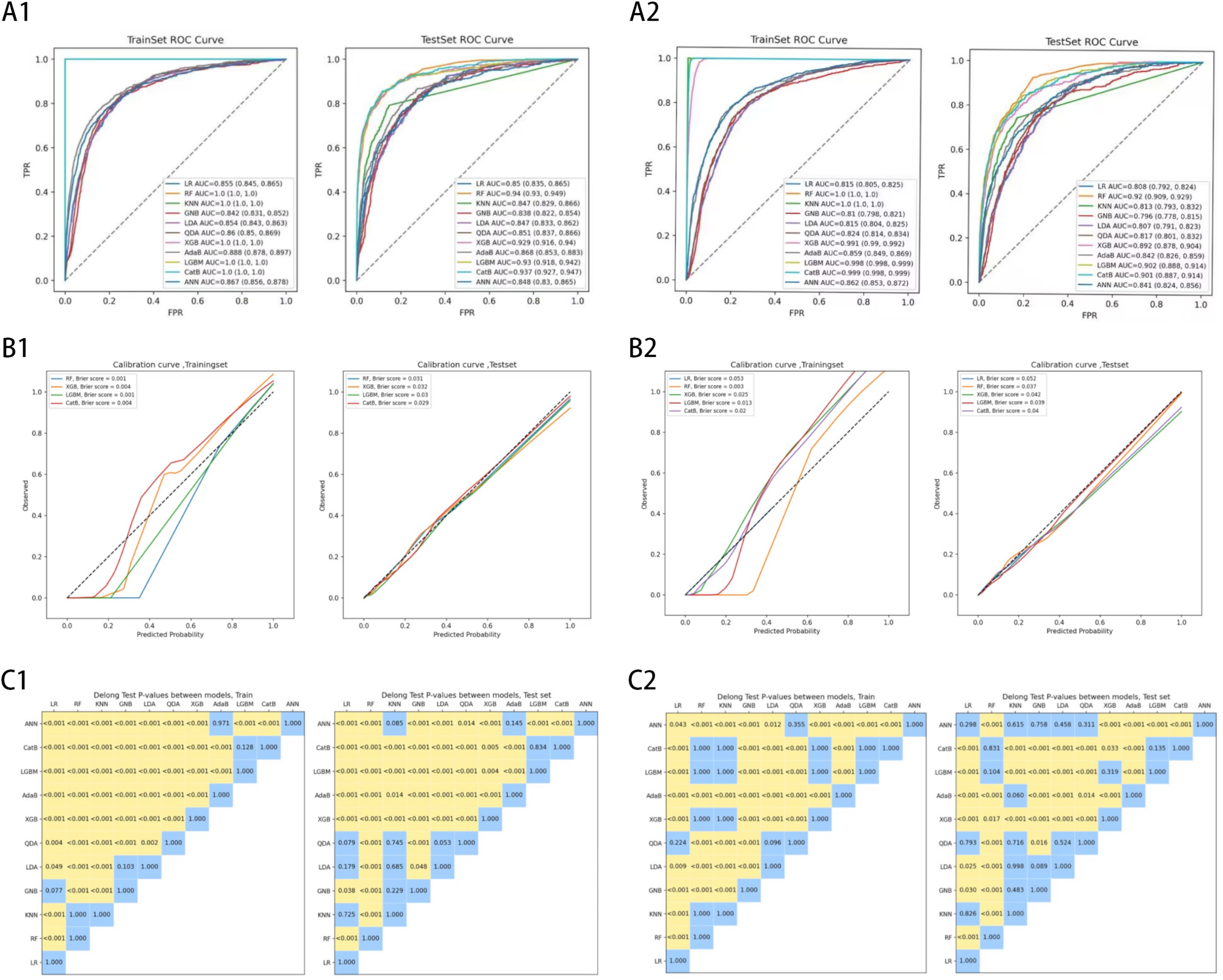

However, further analysis of the confusion matrix revealed that the RF model, while achieving high specificity with a higher prediction risk cutoff threshold, incurred a significantly low sensitivity, indicating a tendency to predict patients as negative. In contrast, the CatBoost model exhibited the highest F1 score on the test set (0.595420), with both Positive Predictive Value (PPV) and Negative Predictive Value (NPV) ranking among the best across all models. The DeLong test comparing the prediction results of the RF and CatBoost models indicated no statistically significant difference in performance (p = 0.500). Calibration curves for the best-performing models also showed that CatBoost achieved the best Brier score of 0.029 on the test set.

### Simplified Model

Figure 4 illustrates the ROC curves for predictive models constructed using Feature Subset B (6 features). Table 5 includes performance evaluation results of Simplified Model. The RF model again achieved the highest AUC of 0.920 on the test set, followed by LightGBM at 0.902, CatBoost at 0.901, and XGBoost at 0.892.

Further analysis of the confusion matrix indicated that the RF model once more used a higher prediction risk cutoff threshold to achieve high specificity, but with very low sensitivity. The CatBoost model maintained the highest F1 score (0.520362) and PPV (0.497835) on the test set. Notably, the DeLong test revealed a statistically significant difference in predictive performance between the RF and CatBoost models (p < 0.001). Calibration curves for the best-performing predictive models indicated that the RF model had the best calibration, achieving the lowest Brier score of 0.037 on the test set, followed by LightGBM at 0.039 and CatBoost at 0.040. While the discriminatory power of the simplified RF model was slightly reduced compared to the full model (0.940 vs. 0.920, p < 0.001), its usability was significantly enhanced.

### Feature Importance and Interpretability Analysis

In the CatBoost model using 13 predictive variables, the most important feature was uric acid, followed by NIHSS score, blood urea nitrogen, and C-reactive protein, along with potassium levels, D-dimer, and age. Figure 5 provides a detailed SHAP value analysis for predicting cerebral herniation outcomes, featuring four distinct panels that collectively offer a comprehensive view of the model’s decision-making process. The bar plot in Figure 5A presents the mean absolute SHAP values for each feature, highlighting their predictive importance. The features bua, nhiss, and urea emerge as the most impactful, with bua exerting a particularly strong influence on the model’s predictions.

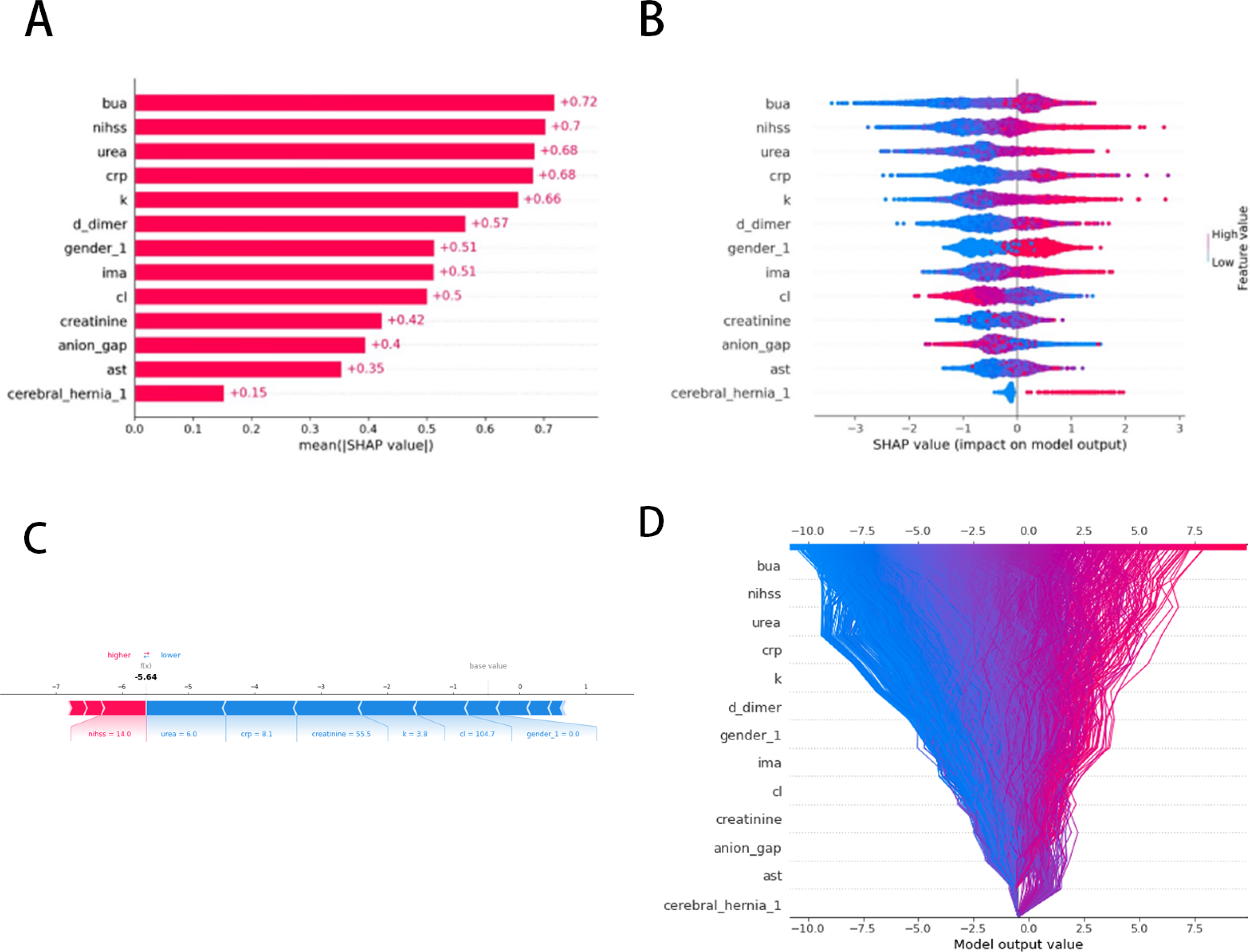

The subsequent dot plot in Figure 5B illustrates the distribution of SHAP values across all samples for each feature, with a color gradient from blue to red indicating the range of feature values. This plot is instrumental for observing the relationship between feature values and their impact on model output, demonstrating how changes in feature values can influence predictions.

The force plot in Figure 5C isolates the prediction for a single sample, delineating the individual contributions of each feature to the model’s output.

The final panel in Figure 5D synthesizes the SHAP values across all samples with a decision plot, mapping these values against the model’s output. The plot’s density visually communicates the concentration of data points, offering a global explanation of the model’s behavior. It reveals the synergistic effect of features on the model’s predictions, underscoring how they collectively shape the outcome.

The trend of SHAP values, mortality risk, and uric acid levels is illustrated in Figure 6A. As uric acid levels increased from 300 μmol/L to 400 μmol/L, the risk of mortality significantly increased, followed by a more gradual change thereafter.

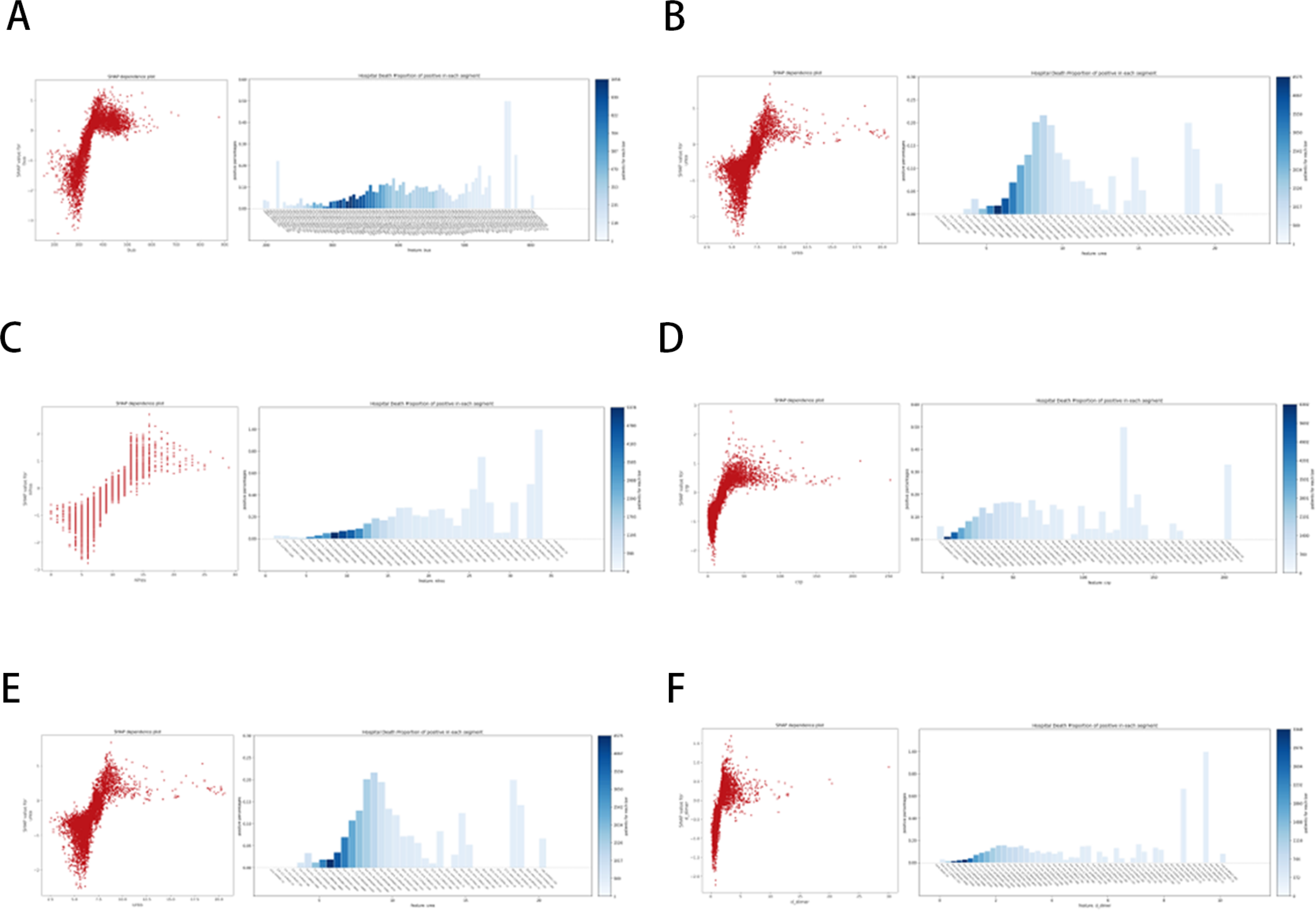

Figure 6B shows the trend of SHAP values, mortality risk, and NIHSS scores, indicating a marked increase in mortality risk with rising NIHSS scores.

Figure 6C illustrates the trend of SHAP values, mortality risk, and blood urea nitrogen levels, where mortality risk increased with blood urea nitrogen levels below 8.5 mmol/L, peaking between 8.5-9.0 mmol/L, followed by a significant decline.

Figure 6D presents the trend of SHAP values, mortality risk, and C-reactive protein levels, where mortality risk consistently increased with rising C-reactive protein levels.

Figure 6E depicts the trend of SHAP values, mortality risk, and D-dimer levels, showing that mortality risk continued to rise with increasing D-dimer levels, stabilizing after reaching 2 mg/L.

Finally, Figure 6F shows the trend of SHAP values, mortality risk, and creatinine levels, where overall, mortality risk significantly increased after creatinine levels exceeded 300 mg/L.

## Discussion

### Summary of Our Study

Stroke is a leading cause of death and disability worldwide, affecting one in four adults over the age of 25 in their lifetime. In China, the situation is even more severe, with an explosive increase in the burden of stroke diseases due to an aging population and the prevalence of unhealthy lifestyles. Ischemic stroke patients account for the majority of stroke cases, making early identification of their mortality risk upon admission and subsequent intervention crucial for improving patient prognosis and reducing disease burden. This study utilized a large sample of patient data from multiple hospitals in Chongqing, comprehensively collecting baseline information such as medical history, laboratory test results, and imaging indicators, to develop a machine learning model for predicting in-hospital mortality. The model consists of two versions; the CatBoost model using thirteen features demonstrated the best predictive performance, while the simplified model using six features, although less effective, is more suitable for emergency situations. Additionally, analysis of related risk factors revealed that uric acid, NIHSS score, blood urea nitrogen, C-reactive protein, and D-dimer significantly impact in-hospital survival.

This study, based on a large sample size and comprehensive clinical, imaging, and laboratory indicators, addressed data quality issues through techniques such as data balancing and imputation. The SHAPRFECV method was employed to select the most predictive feature subset, and the developed model exhibited superior predictive performance compared to previous studies while minimizing the number of required predictive factors for clinical application, enhancing its practical value.

### Comparison with Previous Machine Learning Studies

Many prior studies have established mortality prediction models for ischemic stroke patients using statistical methods, particularly logistic regression, or machine learning approaches. For instance, Bonkhoff et al. predicted in-hospital mortality using machine learning methods based on data from 152,710 ischemic stroke patients, achieving an AUC of 0.90 and a Brier score of 0.109–0.111. Hu et al. utilized data from 5,613 ischemic stroke patients in the MIMIC-IV and eICU-CRD databases, employing an ensemble learning soft voting classifier, yielding AUROC values of 0.861 and 0.844 for internal and external testing, respectively, with AUPRC values of 0.543 and 0.378. However, these studies required 47 and 24 input variables, respectively, complicating clinical application. Matsumoto et al. analyzed data from 4,237 acute ischemic stroke patients using LASSO models to predict all-cause mortality, achieving an AUC of 0.88. However, AUC values for RF and GBDT models were only 0.84, showing no significant advantage over clinical scores like PLAN, IScore, and ASTRAL. The sample size in that study was relatively small, and the predictive performance was inferior to our model, with more than 20 features used, complicating clinical application. Wei Liu et al. developed multiple models based on data from 1,763 ischemic stroke patients using 15 machine learning algorithms. The complete RF model with 58 predictive factors had internal and external validation AUCs of 0.806 and 0.838, respectively; however, performance significantly declined when the feature count was reduced to 35. The RF model using only the top six features resulted in AUCs of 0.795 and 0.830 during internal and external validation, respectively, which may not meet clinical needs. Lea-Pereira et al. established a logistic regression model for 186,245 ischemic stroke patients who did not receive reperfusion therapy, using 10 variables, with training and testing AUCs of 0.742 and 0.736, respectively, indicating moderate predictive capability, while MLP and RF performed even worse. Alejandro Bustamante et al. analyzed data from 12,227 ischemic stroke patients across 42 centers in Spain, using logistic regression and random forests to identify the most significant post-stroke complications affecting in-hospital mortality, but their focus on complications neglected vital laboratory data.

In summary, while several studies have constructed mortality prediction models for ischemic stroke patients, most of these models are highly complex, with insufficient collection of critical indicators, especially laboratory parameters. Additionally, sample sizes and case numbers are relatively small, and predictive performance has not significantly surpassed that of our research. This indicates a need for more scholars to participate in risk assessment for ischemic stroke, and our study provides valuable reference and scientific conclusions.

### Important Feature Analysis

In this study, age and NIHSS score were included as continuous variables. Older patients are likely to have poorer prognoses due to declining physiological function and various underlying diseases. The NIHSS score contributed significantly to the predictive model, ranking second in overall impact in the SHAP analysis, with many individual impacts ranking first, reaffirming its reliability as a predictor of stroke outcomes.

We found that higher uric acid levels were associated with increased in-hospital mortality risk. Previous studies have reported that high uric acid levels increase the risk of stroke; however, whether high uric acid levels serve a protective or harmful role in patient outcomes remains unclear. On one hand, uric acid acts as an effective antioxidant, potentially alleviating oxidative stress after a stroke; on the other hand, elevated uric acid levels may lead to endothelial dysfunction, thrombosis, and arterial occlusion, possibly triggering systemic inflammatory responses. Further large-scale clinical trials are needed to clarify the relationship between uric acid and outcomes in ischemic stroke patients.

In our study, blood urea nitrogen (BUN) levels exhibited a biphasic relationship with mortality risk, peaking between 8.5-9.0 mmol/L. You et al. found that elevated BUN at admission was significantly associated with in-hospital mortality risk among 3,345 ischemic stroke patients in China. Zhou et al. also reported that patients with BUN levels ≥6.74 mmol/L had a significantly increased risk of poor three-month outcomes, although BUN was not identified as an independent risk factor. The interaction between BUN and mortality risk in ischemic stroke patients is complex; low BUN levels may indicate malnutrition, while high levels reflect hemodynamic deterioration, both contributing to poor prognosis. Although our sample size is larger than those studies, the distribution of BUN levels in our cohort was primarily concentrated between 5.0-7.5 mmol/L, where an increasing trend in mortality was evident. Further data collection is needed to clarify these findings.

We also found that higher creatinine levels were associated with increased mortality risk, with average serum creatinine levels significantly higher in deceased patients compared to survivors. High creatinine levels indicate impaired renal function, which has been linked to increased mortality risk in stroke patients. Chronic kidney disease can worsen vascular dysfunction, calcification, and arteriosclerosis, all of which elevate stroke risk. Furthermore, renal dysfunction critically impacts recovery post-stroke. While contrast angiography can aid in diagnosing and treating acute stroke, iodinated contrast agents may further impair renal function, leading to poorer outcomes.

Additionally, we identified elevated potassium levels as a risk factor for in-hospital mortality. Huang et al. reported higher median potassium levels in deceased patients among 1,236 elderly ICU patients with ischemic stroke. Mittal et al. also noted higher potassium concentrations in patients who died within 30 days post-admission. Longitudinal studies have found a positive correlation between serum potassium levels and mortality rates. However, some studies indicate that low potassium levels are also associated with poor functional outcomes and mortality in ischemic stroke patients, possibly due to elevated catecholamine levels leading to potassium uptake by cells. Considering the poor prognosis of patients with renal impairment, we hypothesize that elevated potassium levels may result from excessive potassium release due to cell death, compounded by renal dysfunction, arrhythmias, and electrolyte imbalances.

Our study found that elevated D-dimer levels were associated with increased mortality risk. D-dimer is a degradation product of fibrin and indicates a hypercoagulable state. Previous studies have shown that high D-dimer levels increase the risk of ischemic stroke. Additionally, D-dimer may be linked to the activation of inflammatory responses, and our findings further support its role as a biomarker for ischemic stroke prognosis.

Previous research has indicated that C-reactive protein (CRP) reflects the degree of inflammation caused by cerebral infarction and can be used to predict outcomes in ischemic stroke patients. CRP is a glycoprotein produced by the liver that can be rapidly upregulated by inflammatory cytokines, exacerbating ischemic brain injury. Furthermore, CRP may promote the development and progression of atherosclerosis, activate the complement system, inhibit fibrinolysis, and facilitate thrombosis, thereby increasing neuronal damage. Elevated CRP levels likely indicate greater brain necrosis and more severe stroke.

### Limitations and Future Directions

Larger Multicenter Validation and Increased Sample Size While this study’s sample size is advantageous compared to previous research, larger samples can more accurately capture real-world patterns for machine learning models. Future studies should consider expanding the sample size to enhance the reliability and generalizability of the model. This study included only patients from the Chongqing region, and results may be influenced by local lifestyle factors. To develop a more universally applicable predictive model, it is crucial to include patients from more diverse geographical areas for validation and adjustment. Attempts to utilize large open-source databases like MIMIC-IV and eICU-CRD were unsuccessful due to their non-specific focus on stroke populations and significant missing variables such as NIHSS scores.

Incorporating Causal Inference to Analyze Deep Interconnections Among Features

This study employed interpretability methods to demonstrate the contributions of variables in the model but could be further enhanced by causal inference methods to analyze relationships among variables, including mediating effects and confounders. This would deepen clinicians’ understanding of the model and improve trust in its application.

Consideration of Post-Baseline Features

This study developed an early model based on admission characteristics. Although standardized treatments were used, future research should consider the impact of post-baseline features on patient outcomes to enhance the model’s practicality.

In conclusion, while this study achieved significant results, further improvements are needed in sample size, data sources, and variable analysis.

## Methods

### Patient Cohort

This study retrospectively collected data from patients with ischemic stroke admitted between June 2017 and July 2023 at the Chongqing Emergency Medical Center, Yubei District Traditional Chinese Medicine Hospital, Qianjiang Central Hospital, and Bishan District People’ s Hospital.

#### Inclusion Criteria

Age 18 years or older.

Diagnosis of ischemic stroke upon admission (International Classification of Diseases, ICD-10, I63) (including cerebral infarction, lacunar infarction, and old cerebral infarction).

Treatment followed the guidelines outlined in the “Chinese Guidelines for the Diagnosis and Treatment of Acute Ischemic Stroke” and the “Consensus of Chinese Experts on Emergency Management of Acute Ischemic Stroke.”

#### Exclusion Criteria

Patients with other potential causes of in-hospital mortality or concurrent diseases, such as advanced malignant tumors, severe traumatic brain injury (TBI), severe autoimmune diseases, etc., were excluded.

The patients were randomly divided into training and testing groups in a 7:3 ratio. The overall inclusion process and data handling steps are illustrated in Figure 1.

### Data Collection and Preprocessing

Baseline clinical information was collected, including demographics (sex, age), medical history and comorbidities (diabetes, hypertension, coronary heart disease, fatty liver, etc.), laboratory test results (complete blood count, liver function, kidney function, cardiac enzyme levels, etc.), and imaging indicators (e.g., extent of brain lobe involvement determined by CT or MRI and vascular stenosis determined by CTA, DSA, or MRA). The NIHSS score was assigned by a neurosurgical specialist based on the patient’s clinical course upon admission to assess the severity of the stroke. Subsequently, the in-hospital survival status and cause of death were determined from patient records.

### Data Imputation

Variables with more than 10% missing values and can’t be remedy by doctors were excluded. Remaining variables were imputed using the missForest method, an iterative imputation approach based on random forests, which has been validated in prior studies. During the imputation process, patient outcomes were not utilized to prevent data leakage.

Data Balancing

Given the high imbalance in the in-hospital mortality/survival ratio (with an in-hospital mortality rate of only 6%), various methods were employed to balance the dataset, including SMOTE, SMOTE-omek, ADASYN, random oversampling, and random undersampling. The effectiveness of these methods was assessed by training a random forest model on the balanced data using five-fold cross-validation.

### Feature Reduction

To reduce model complexity, the SHAPRFECV method was employed to select the optimal feature subset. This tree-based binary classification algorithm uses feature importance scores derived from SHAP analysis as a reference for Recursive Feature Elimination (RFE), ensuring high efficiency. Additionally, the variance inflation factor (VIF) was calculated to confirm that the selected feature set did not exhibit severe multicollinearity. For model development, we selected two feature subsets of differing sizes to create models of varying complexity and generalizability.

### Model Development

Using the selected feature subsets, multiple machine learning models were established to predict in-hospital mortality. The specific models included: Logistic Regression (LR), Random Forest (RF), K-Nearest Neighbors (KNN), Gaussian Naive Bayes (GNB), Linear Discriminant Analysis (LDA), Quadratic Discriminant Analysis (QDA), Extreme Gradient Boosting (XGBoost), Adaptive Boosting (AdaBoost), Light Gradient Boosting Machine (LightGBM), CatBoost, and Artificial Neural Networks (ANN). Each model underwent three-fold cross-validation and grid search to determine the optimal hyperparameters, which were then applied to fit the entire training set. The performance of each model was subsequently evaluated on the test set.

To assess model calibration, isotonic regression was used to adjust the predicted probabilities from each model.

### SHAP Analysis

To further analyze the contribution of each feature in the model, SHAP (SHapley Additive exPlanations) values were employed to interpret the variables in the final model.

## Statistical Methods and Libraries

PostgreSQL v15 (http://www.postgresql.org/) was utilized to search and extract data from the local database. The open-source Python library SciPy (v1.7.3) was employed for statistical analysis, with the missForest method sourced from the missingpy package (v0.2.0). The SHAPRFECV method was obtained from the probatus package (v1.8.9). The CatBoost model was implemented using the catboost package (v1.2.3), the LightGBM model from the lightgbm package (v4.3.0), and the XGBoost model from the xgboost package (v1.6.2), while the remaining machine learning models were sourced from scikit-learn (v1.0.2).

The maximum Youden index was calculated from the ROC curve of the training set for each model to determine the optimal classification threshold, which was then applied to the same models on the test set. Model discrimination was assessed using the receiver operating characteristic (ROC) curve and the area under the curve (AUC). Model calibration was evaluated with calibration curves and Brier scores, while the clinical decision curve analysis (DCA) measured the net benefits of the model across different thresholds. AUC values and their confidence intervals from the five-fold cross-validation in the training set were calculated using the R package cvAUC, while confidence intervals for other AUC values and Brier scores were obtained through bootstrapping with 1,000 resamples on the respective datasets.

The differences between the prediction results of the models were assessed using the Fast DeLong test, an efficient version of the DeLong test, as described in the WROC package (v3.6.3). All binary classification thresholds for predicted probabilities from the models were established using the maximum Youden index from the training cohort. Throughout the study, a two-tailed p-value of <0.05 was considered statistically significant.

### Conclusion

This study utilized large sample patient data from four hospitals in Chongqing, comprehensively incorporating patients’ historical features, laboratory test results, and imaging indicators to develop a machine learning model based on baseline characteristics for predicting in-hospital mortality. By maximizing model simplicity, we enhanced its convenience for clinical use while ensuring predictive efficacy. Additionally, the study conducted an in-depth analysis of related risk factors, providing important references for clinical decision-making. The development of this model not only aids in improving clinical workflow efficiency but also establishes a foundation for further research and optimization in the management of stroke patients. Future validation studies involving more diverse regions and larger sample sizes will further enhance the model’s generalizability and applicability.

## Data Availability

All data can be available from

## Author Contributions

JL and HH are both first writer who analysed the data by python and wrote the first draft of the manuscript.YD and XJ are both corresponding author who wrote part of the draft and designed the original research. Chongqing University Central Hospital,Chongqing University Qianjiang Hospital, Yubei District hospital and Bishan hospital of Chongqing Medical University provided the database of all cases of the patients. The others collected data and wrote sections of the manuscript. All authors took part in the research and contributed to manuscript revision, read,and approved the submitted version.

## Data availability statement

The codes,models,analysis results will be uploaded at https://github.com/conanan. The full dataset can be provided for researchers if needed by the corresponding author.

## Acknowledgements

The authors would like to thank the colleagues in the information and imaging departments for their hard work contributing to the final research results.

## Ethics approval statement

We confirm that we have read the Journal’s position on issues involved in ethical publication and affirm that this report is consistent with those guidelines.

## Patient consent statement

This study was a retrospective study and only deidentified patient data were collected, exempting the need for patient informed consent rights.

## Permission to reproduce material from other sources

There are no reproduce material from other sources.

## Clinical trial registration

The trail number is RS202406.

